# Reporting of medication adherence in randomized controlled trials of pharmacological interventions for SARS-CoV-2: a cross-sectional analysis

**DOI:** 10.1101/2022.10.30.22281709

**Authors:** C K Lee, A Otunla, Brennan, JK Aronson, D Nunan

## Abstract

**Background:** Adherence to pharmacological interventions in clinical trials is crucial for correct estimation of beneficial and adverse effects, including trials of SARS-CoV-2. The Template for Intervention Description and Replication (TIDieR) – a 12-item extension of the Consolidated Standards of Reporting Trials (CONSORT) reporting guidelines – includes two items (11 and 12) that address intervention adherence reporting in trial publications.

**Objective:** To assess compliance with TIDieR items 11 and 12 of randomised controlled trials (RCTs) of interventions in SARS-CoV-2 infection published in five selected journals during 2021.

**Methods:** We assessed SARS-CoV-2 pharmacological RCTs published in the *Annals of Internal Medicine, The BMJ, JAMA, The Lancet*, and *The New England Journal of Medicine* in 2021 for compliance with TIDieR items 11 and 12. Item 11 was assessed in two parts: 11a—how intervention adherence was assessed; 11b—if any strategies were used to maintain or improve how intervention adherence was maintained or improved. Item 12 assessed the extent to which the intervention was delivered as planned. We calculated raw adherence and proportional (weighted) adherence for pharmacological and comparator interventions where available.

**Results:** We found 75 eligible RCTs, of which 28 (37%) reported results related to SARS-CoV-2 vaccinations. Compliance with items 11a and 12 could be assessed in 71 of these 75. Of those 71 RCTs, 37 (52%, 95% confidence interval 40–64%) were compliant with reporting of item 11a. Seven RCTs had a strategy to assess compliance with item 11b, and only three (43%, 9–82%) of those complied with item 11b reporting. Of the 71 RCTs, 70 complied with reporting of item 12. Only one of the 71 RCTs (1.4%, 0–7.6%) fully complied with TIDieR items 11a, 11b, and 12. Compliance varied across journals.

**Conclusions:** RCTs of SARS-CoV-2 pharmacological interventions published in high-impact medical journals complied variably with reporting of intervention adherence, even though the journals endorse CONSORT. The implications for interpretation, application, and replication of findings based on these publications warrant consideration.

## Introduction

Many different terms have been used interchangeably and inexactly to describe adherence to pharmacological interventions.^1–3^ To resolve this unclear taxonomic structure,^4^ the Ascertaining Barriers to Compliance (ABC) project published a systematic literature search on the evolution of terms related to medication-taking behaviour. The ABC authors then proposed a new set of terms and a related taxonomy defining medication adherence nomenclature,^5^ which has since been widely used. The ABC authors defined medication adherence as a process comprising three steps: initiation, implementation (the extent to which the medication given corresponds with the prescribed protocol), and discontinuation. This multi-step definition accounts for all aspects of pharmacological intervention adherence and highlights the multiple factors that contribute to intervention delivery.

Adherence to pharmacological therapy, in both research and clinical settings, is crucial for accurately ascertaining the benefits and harms of such therapy.^4,6^ In research settings, variable adherence to study intervention(s) is a mediating factor that can significantly bias results and reduce study power.^7^ In the case of pharmacological interventions, poor intervention adherence can result in underestimation of drug efficacy and adverse effects, impaired development of breakthrough drugs and drugs for rare diseases, and ultimately treatment failures.^8–10^ Reporting of adherence to pharmacological interventions is variable across clinical trials.^10^

The Consolidated Standards of Reporting Trials, 2010 (CONSORT) guidelines^11^ is a consensus-based set of recommendations that provides suggested minimum reporting requirements for randomised trials and aims to encourage accurate scientific reporting.^12^ CONSORT statement 5 recommends the use of the Template for Intervention Description and Replication (TIDieR) extension^12^ to ensure good reporting of trial interventions. TIDieR extension checklist items 11 and 12 specifically address adherence to trial interventions (Box 1). Several biomedical journals require or recommend the use of CONSORT, and therefore the TIDieR checklist, as part of their publishing process.

During 2021, publication of trials of interventions for preventing and treating SARs-CoV-2 infection was streamlined.^13–16^ It is important to maintain reporting standards in order to communicate scientific rigour and the effects of interventions, particularly during a public health crisis. Here we assess the reporting standards of adherence to interventions in randomised control trials related to SARS-CoV2 published in five major medical journals during 2021.

### Box 1.

TIDieR checklist items 11 and 12

11. Planned: If intervention adherence or fidelity was assessed, describe how and by whom, and if any strategies were used to maintain or improve fidelity, describe them.

12. Actual: If intervention adherence or fidelity was assessed, describe the extent to which the intervention was delivered as planned.

[Note: The TIDieR guideline explanation document gives the following definition of “fidelity”: the degree to which an intervention happened in the way the investigators intended it to.^17^]

## Methods

We performed a cross-sectional study of RCTs published in five leading medical journals: *Annals of Internal Medicine* (*Ann Intern Med*), *The BMJ, JAMA, The Lancet*, and *The New England Journal of Medicine* (*NEJM*). We selected these journals because they have some of the highest impact factors among general medical journals, with wide readerships, and are highly influential among health professionals, health journalists, and policy makers.

We conducted a search through PubMed for RCTs assessing pharmacological interventions containing the keywords SARS* OR Covid*, published between 01/01/2021 and 31/12/21. We filtered these results using the Journal filter (selecting the five aforementioned journals) and the RCT filter in PubMed.

### Inclusion criteria

A publication was included in the analysis if it met the following criteria: (1) the RCT was published between 01/01/2021 and 31/12/21; (2) the RCT was published in one of the five aforementioned journals in any format, including letters to the editor; (3) the RCT investigated a pharmacological intervention related to the prevention or treatment of SAR-CoV-2. This was defined as any drug that had primary, secondary, and/or tertiary drug preventive effects. Oxygen therapy and oxygen therapy delivery methods were also included, as oxygen is a prescribed pharmacological therapy in UK healthcare.

If trials did not meet all of the above criteria, they were excluded from the analysis.

### Data extraction and assessment of reporting compliance with the TIDieR guidelines

The RCTs that met the above inclusion criteria underwent title and abstract screening in duplicate by CL and DN. Discrepancies were resolved by discussion. Using a standardised form, the following information was extracted in duplicate (CL, AO, or IB) from each included trial: publishing journal, authors, PubMed identification number, article title, type of intervention, and intervention drug class. Drug class was recorded as defined in the article. When intervention adherence was published, both investigated pharmacological intervention and control intervention adherence data were recorded. Adherence rate was calculated from the raw data using the population randomized to the intervention and the number of participants receiving the pharmacological intervention, as detailed in the trial protocol.

Data extraction to assess article compliance with TIDieR guideline items 11 and 12 was performed independently (CL), and a subset (25% of included articles) was screened in duplicate (DN); discrepancies were resolved by discussion. TIDieR item 11 has multiple requirements, and so compliance with that item was assessed in two parts:

11a. Planned: If intervention adherence or fidelity was assessed describe how and by whom.

11b. Planned: If intervention adherence or fidelity was assessed, and if strategies were used to maintain or improve fidelity, describe them.

Item 12 was assessed as published:

12. Actual: If intervention adherence or fidelity was assessed, describe the extent to which the intervention was delivered as planned.

Items 11a, 11b, and 12 contain the conditional requirement ‘if intervention adherence or fidelity was assessed’. Compliance with reporting guidelines was assessed only in trials in which this conditional requirement was met. If intervention adherence or fidelity was not assessed, the trial was scored as “not applicable” for that item. The outcome for each item could therefore be: compliant, noncompliant, or not applicable. Data were entered into an Excel spreadsheet, and proportions with 95% confidence intervals (CI) and overall weighted proportions were calculated.

The extent to which the TIDieR checklist is endorsed by the journals included was assessed from the instructions to authors regarding TIDieR or CONSORT guidelines (Table 1). We categorised journals as requiring TIDieR if the author instructions included the terms ‘must’, ‘required’, or ‘should’ and as recommending the use of the TIDieR or CONSORT guidelines if the instructions to authors included the terms “should”, “recommend”, “follow”, or “encourage”.

**Table 1.** Journal endorsement of TIDieR or CONSORT guidelines

| Degree of TIDieR or CONSORT endorsement | Journal | Author instruction |
| --- | --- | --- |
| Recommended | Ann Intern Med | ‘Follow relevant reporting recommendations. The EQUATOR site includes the following guidelines, which *Annals* endorses: Controlled Trials: CONSORT’<sup>18</sup> |
|  | NEJM | ‘Authors should provide a flow diagram in CONSORT format. The editors also encourage authors to submit all the relevant information include in the CONSORT checklist. Although all of this information may not be published with the manuscript, it should be provided in either the manuscript or a supplementary appendix at the time of submission’<sup>19</sup> |
| Required | The Lancet | ‘Reports of trials must conform to CONSORT 2010 guidelines and should be submitted with their protocols’ ‘Cluster-randomised trials must be reported according to CONSORT extended guidelines’ ‘Randomised trials that report harms must be described according to extended CONSORT guidelines’<sup>20</sup> |
| Checklist | BMJ | ‘For a [sic] clinical trials, use the CONSORT checklist and also include a structured abstract that follows the CONSORT statement for abstract checklist, the CONSORT flow chart, and where applicable, the appropriate CONSORT extension statements (for example, for cluster RCTs, pragmatic trials, etc.’<sup>21</sup> |
|  | JAMA | ‘Manuscripts reporting the results of randomized trials must include the CONSORT flow diagram showing the progress of patients throughout the trial. The CONSORT checklist should also be completed and submitted with the manuscript’<sup>22</sup> |

## Results

### Sample demographics

We identified 167 articles in our initial search, of which 75 (45%) were eligible for inclusion (Figure 1).

**Figure 1.**
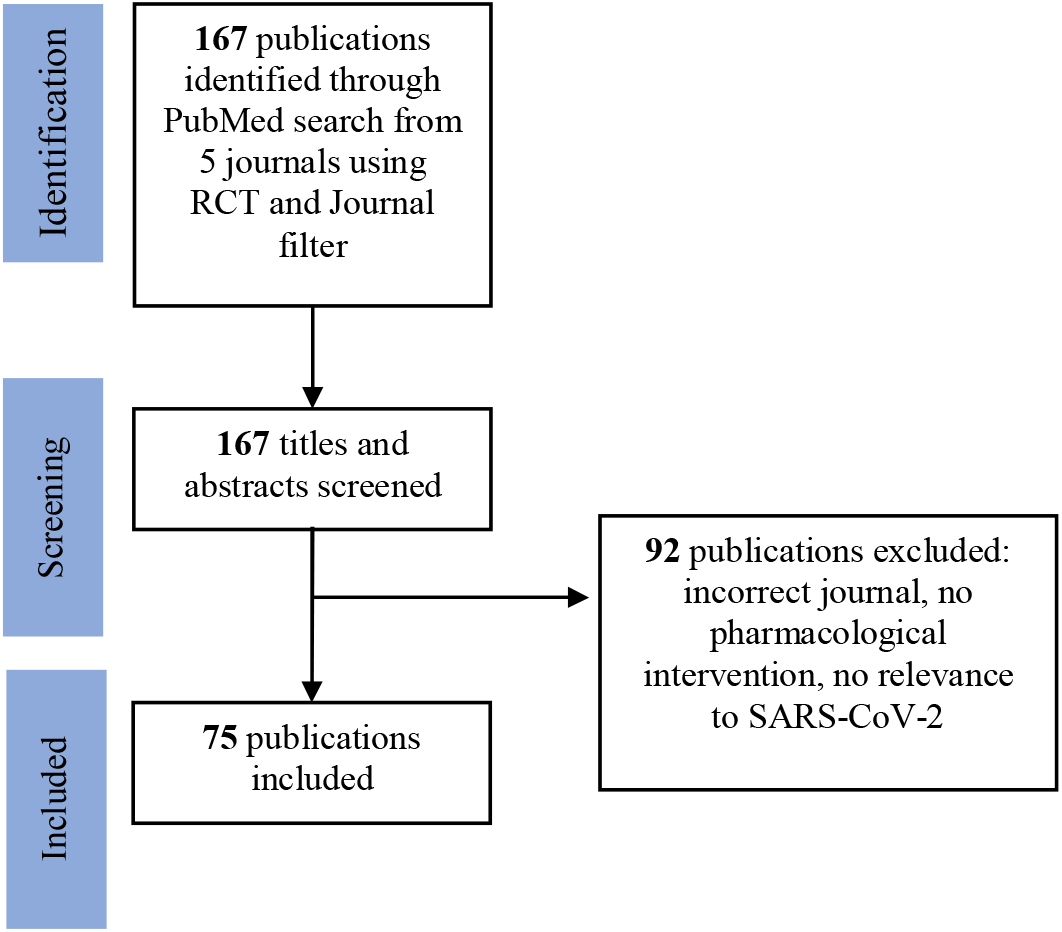
Flow chart of the search

Table 2 shows the distribution of articles included in the final sample group. Most of the articles included were published in *NEJM* and *The Lancet*.

**Table 2.** Articles included per journal

| Journal | No. of articles in final sample |
| --- | --- |
| Ann Intern Med | 2 |
| BMJ | 3 |
| JAMA | 15 |
| The Lancet | 21 |
| NEJM | 34 |

The final sample included a broad range of pharmacological interventions studied, including vaccination, antimalarial therapies, and anticoagulants (Table 3).

**Table 3.** Pharmacological interventions studied in included articles

| **Intervention drug class** | **Number of trials included** |
| --- | --- |
| Vaccine | 28 |
| Monoclonal antibody | 15 |
| Anticoagulant | 7 |
| Convalescent plasma | 5 |
| Corticosteroid | 4 |
| Antimalarial drug | 3 |
| Macrolide antibiotic | 3 |
| Janus kinase inhibitor | 2 |
| Oxygen | 2 |
| Antiplatelet drug | 1 |
| Antiparasitic drug | 1 |
| ACE inhibitor | 1 |
| Vitamin D supplementation | 1 |

### TIDieR compliance

TIDieR items 11a, 11b, and 12 were “not applicable” to 4/75 articles (5.3%, 1.5–13%). Overall, 37/71 (52%, 40–64%) articles were compliant with item 11a.

Only seven articles had a fidelity strategy and could be assessed for compliance with 11b. Three of those seven studies (43%, 9.0–82%) with a fidelity strategy complied with item 11b reporting standards.

Of the 71 articles, 70 (99.0%, 92.4–100%) complied with TIDieR item 12.

Thus, 1/71 (1.4%, 0–7.6%) articles fully complied with TIDieR items 11a, 11b, and 12. Compliance and weighted compliance varied across journals (Table 4).

**Table 4.**
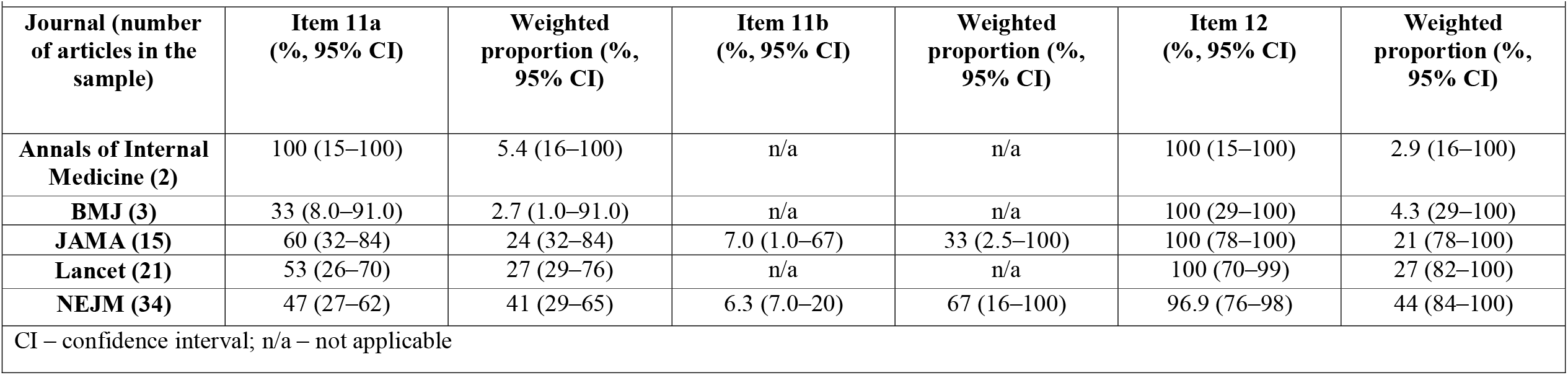
Compliance with TIDieR checklist items 11a, 11b, and 12

### Intervention adherence

Of the 75 articles included in our sample, 71 reported adherence to the intervention and placebo. Mean sample intervention adherence was reported as 92.3% (88–96.2%) and placebo adherence 90.3% (86–95.0%). In vaccine trials (25/71), adherence to both intervention (97.8%, 96.8–98.9%) and placebo (97.5%, 96.3–98.7%) injections was higher than adherence in non-vaccine intervention trials (46/71): intervention 91.4% (87–95.9%), placebo 87% (81–94.2%).

## Discussion

### Summary of main findings

We have examined the reporting standards of adherence to trial interventions in coronavirus-related publications from five major academic journals during 2021, using the TIDieR checklist items 11 and 12. There was poor compliance with item 11a (37/71, 52%, 40–64%). There was little applicability of item 11b (7/71, 9.9%, 4.1–19%), and of these seven articles, three (43%, 9.0–82%) complied with item 11b. We found good compliance with TIDieR item 12 across the sample set (70/71, 99.0%, 92.4–100%).

CONSORT (2010) Item 5 recommends that the TIDieR checklist should be used in conjunction with CONSORT guidelines in randomised controlled trials. Journals that require authors to adhere to CONSORT surprisingly had poorer compliance with reporting for TIDieR items 11a and 12. *The Lancet* requires CONSORT guidelines to be adhered to, but had a low weighted compliance proportion with item 11a (27%) and item 12 (27%). Similarly, *BMJ* (2.7% and 4.3%) and *JAMA* (24% and 21%) both had low weighted compliance proportions for items 11a and 12 even though both journals *require* a completed CONSORT checklist as part of manuscript submission. In contrast, adherence to items 11a (41%) and 12 (44%) was higher in the *NEJM*, which only *recommends* the use of CONSORT.

Raw adherence to 11a was modestly higher in *JAMA* and *The Lancet* than in other journals. Only the *NEJM* did not meet item 12 in all articles (96.9%).

Four of the 75 trials in our sample did not report intervention adherence. Of these four, one was published as a “Letter to the Editor” and three were published as full manuscripts. All four articles assessed single-dose interventions administered by healthcare professionals (three assessed vaccines and one studied subcutaneous delivery of a monoclonal antibody). The Letter and one of the three papers published in full did not acknowledge intervention adherence. Two of the three articles (in *NEJM* and *The Lancet*) published in full, acknowledged intervention in their protocol, stating that compliance was not relevant, as the intervention was administered by research clinical staff and not by participants. However, it is wrong to assume that administration of an intervention by a skilled worker in a clinical or trial setting necessarily guarantees adherence. Although it removes some common factors that reduce adherence, namely, patient behaviour, education, and skill,^23–25^ other factors affecting intervention adherence are introduced, such as patient attendance, work environment, and provider skill. Broadly, given the ABC multi-step definition of adherence, to ensure that the correct intervention protocol is delivered, intervention adherence should always be considered in study design and reporting.

We have recorded overall higher adherence rates in vaccination trials than in non-vaccine trials in our sample. This included trials with multiple vaccinations. This may be because those conducting such trials are keenly aware of the importance of adherence, but selection bias of participants for primary prevention trials might also account for it.

Despite 34/71 (48%) noncompliance with item 11a (describing the methods used to assess adherence), we found 99% compliance with TIDieR reporting standard item 12, by which articles must describe the extent to which the trial intervention was delivered as planned. In order to calculate intervention adherence, there must be a well described method of assessment of adherence. Thus, the discrepancy between compliance with items 11a and 12 demonstrates poor reporting across all journals, except the *Annals of Internal Medicine*.

### Implications

Despite the pursuit of efficacious interventions related to SARS-CoV-2 infection, few articles in our sample detailed strategies to ensure that interventions were delivered per protocol. Overall, poor compliance with item 11, compared with 99% reported compliance with item 12, shows the need for increased emphasis on reporting adherence methods. We have examined the standards of reporting, not the scientific rigour nor the quality of the data of the trials included in our sample, and no conclusions about benefits or harms of the interventions may be extrapolated from our assessment of reporting standards.

### Limitations

Our sample included articles from five medical journals during 2021, and so conclusions about reporting standards cannot be extended beyond these journals and time frame.

Most of the articles we included were published by *NEJM, The Lancet*, and *JAMA. The BMJ* (n=3) and *Annals of Internal Medicine* (n=2) contributed only a few articles. We compensated for this by calculating both raw and weighted proportions. A more accurate assessment of reporting standards might be provided by analysing equal number of articles from each journal, although doing so would require markedly widening the time frame of publication.

The degree of TIDieR endorsement is extrapolated from journal endorsement of the CONSORT (2010) guidelines and checklist. CONSORT item 5 recommends the use of the TIDieR checklist as an extension to the CONSORT guidelines. However, the journals that recommend CONSORT in their publication process only weakly endorse the TIDieR checklist.

## Conclusions

Most of the RCTs of SARS-CoV-2 pharmacological interventions published in five leading medical journals during 2021 did not comply with TIDieR checklist items 11 and 12. Future work should address compliance with TIDieR items 11 and 12, through discussion with authors and journals.

## Data Availability

The repository of extracted data is available on the Open Science Framework.

https://osf.io/b96g3/files/osfstorage/635e587e0fac8308f979c6df

https://osf.io/b96g3/files/osfstorage/635e587eb233120900430b40

## Declarations

### Statement of ethical approval

We have used publicly available information, for which ethics committee approval is not required.

### Contributorship statement

JKA suggested the idea for this study, which was built upon by DN; both wrote the initial study protocol. This protocol was later edited by CL in collaboration with JKA and DN. CL, AO, IB, and DN contributed to data extraction. CL analysed extracted data, with advice and direction from JKA and DN. CL, JKA, and DN contributed to authorship of this manuscript and all authors approved the final version.

### Data sharing

Data are extracted from publicly available manuscripts. The source data were openly available before initiation of this study. A list of manuscripts from which data were extracted is available here:

https://osf.io/b96g3/files/osfstorage/635e587e0fac8308f979c6df

https://osf.io/b96g3/files/osfstorage/635e587eb233120900430b40

The data can be found by searching for these studies through PubMed, where each study is openly available:

The repository of extracted data is available on the Open Science Framework:

https://osf.io/b96g3/files/osfstorage/635e587e0fac8308f979c6df

https://osf.io/b96g3/files/osfstorage/635e587eb233120900430b40

## References

1. Apt L, Barrett AB. Patient compliance vs cooperation. Arch Ophthalmol 1987 Mar;105(3):315.

2. Lehane E, McCarthy G. Medication non-adherence--exploring the conceptual mire. Int J Nurs Pract 2009 Feb;15(1):25–31.

3. Romano PE. Semantic follow-up: adherence is a better term than compliance is a better term than cooperation. Arch Ophthalmol 1988 Apr;106(4):450.

4. Steiner JF, Earnest MA. The language of medication-taking. Ann Intern Med 2000 Jun 6;132(11):926–30.

5. Vrijens B, De Geest S, Hughes DA, Przemyslaw K, Demonceau J, Ruppar T, Dobbels F, Fargher E, Morrison V, Lewek P, Matyjaszczyk M, Mshelia C, Clyne W, Aronson JK, Urquhart J; ABC Project Team. A new taxonomy for describing and defining adherence to medications. Br J Clin Pharmacol 2012 May;73(5):691–705.

6. Cals JWL. Comment on: The higher the number of daily doses of antibiotic treatment in lower respiratory tract infection the worse the compliance. J Antimicrob Chemother 2009 May;63(5):1083–4; author reply 1084-5.

7. Brown MT, Bussell JK. Medication adherence: WHO cares? Mayo Clin Proc 2011 Apr; 86(4): 304–14. Available from: https://pubmed.ncbi.nlm.nih.gov/21389250.

8. Janjua S, Pike KC, Carr R, Coles A, Fortescue R, Batavia M. Interventions to improve adherence to pharmacological therapy for chronic obstructive pulmonary disease (COPD). Cochrane Database of Systematic Reviews 2021 Sep 8;9(9):CD013381. Available from: https://www.cochranelibrary.com/cdsr/doi/10.1002/14651858.CD013381.pub2/full.

9. Kini V, Ho PM. Interventions to improve medication adherence: a review. JAMA 2018 Dec 18;320(23):2461–73.

10. Breckenridge A, Aronson JK, Blaschke TF, Hartman D, Peck CC, Vrijens B. Poor medication adherence in clinical trials: consequences and solutions. Nat Rev Drug Discov 2017 Mar;16(3):149–50.

11. Conn VS, Ruppar TM. Medication adherence outcomes of 771 intervention trials: systematic review and meta-analysis. Prev Med 2017 Jun;99:269–76.

12. Hoffmann TC, Glasziou PP, Boutron I, Milne R, Perera R, Moher D, Altman DG, Barbour V, Macdonald H, Johnston M, Lamb SE, Dixon-Woods M, McCulloch P, Wyatt JC, Chan AW, Michie S. Better reporting of interventions: template for intervention description and replication (TIDieR) checklist and guide. BMJ 2014 Mar 7;348:g1687.

13. Škorić L, Glasnović A, Petrak J. A publishing pandemic during the COVID-19 pandemic: how challenging can it become? Croat Med J 2020 Apr;61(2):79–81.

14. Wolkewitz M, Puljak L. Methodological challenges of analysing COVID-19 data during the pandemic. BMC Med Res Methodol 2020 Apr 14;20(1):81.

15. Peyrin-Biroulet L. Will the quality of research remain the same during the COVID-19 pandemic? Clin Gastroenterol Hepatol 2020 Aug;18(9):2142.

16. Majumder MS, Mandl KD. Early in the epidemic: impact of preprints on global discourse about COVID-19 transmissibility. Lancet Glob Health 2020 May;8(5):e627–30.

17. Carroll C, Patterson M, Wood S, Booth A, Rick J, Balain S. A conceptual framework for implementation fidelity. Implement Sci 2007 Nov 30;2:40.

18. Annals of Internal Medicine. Information for Authors. https://annals.org/aim/pages/authors. Accessed June 16, 2022.

19. New England Journal of Medicine. Information for Authors. https://www.nejm.org/doi/full/10.1056/NEJM197001012820120. Accessed June 16, 2022.

20. The Lancet. Information for Authors. https://www.thelancet.com/pb/assets/raw/Lancet/authors/tlgh-info-for-authors.pdf. Accessed June 16, 2022.

21. JAMA Network. Instructions for Authors. https://jamanetwork.com/journals/jama/pages/instructions-for-authors. Accessed June 16, 2022.

22. BMJ. Article types and preparation. https://www.bmj.com/about-bmj/resourcesauthors/article-types. Accessed June 16, 2022.

23. Fisher JD, Fisher WA. Changing AIDS-risk behavior. Psychol Bull 1992 May;111(3):455–74.

24. Yang C, Hui Z, Zeng D, Liu L, Lee DTF. Examining and adapting the information-motivation-behavioural skills model of medication adherence among community-dwelling older patients with multimorbidity: protocol for a cross-sectional study. BMJ Open 2020 Mar 24;10(3):e033431.

25. Kvarnström K, Westerholm A, Airaksinen M, Liira H. Factors contributing to medication adherence in patients with a chronic condition: a scoping review of qualitative research. Pharmaceutics 2021 Jul 20;13(7):1100.

